# High rate of increased level of plasma Angiotensin II and its gender difference in COVID-19: an analysis of 55 hospitalized patients with COVID-19 in a single hospital, WuHan, China

**DOI:** 10.1101/2020.04.27.20080432

**Authors:** Na Liu, Yan Hong, Ren-Gui Chen, Heng-Mei Zhu

**Author notes:** Correspondence to Heng-Mei Zhu. Na Liu and Yan Hong contributed to the work equally. Funding Statement: This study was funded by Basic Research Project of Shenzhen Science and Technology Innovation Commission (JCYJ20160429181842402).

## Abstract

**Background:** 2019 Novel coronavirus disease (COVID-19) is turning into a pandemic globally lately. Angiotensin-converting enzyme 2 (ACE2) is identified as an important functional receptor for SARS-Cov-2. ACE2 and ACE are homologues with inverse functions in the renin–angiotensin system. ACE converts angiotensin I into a vital vasoactive peptide called angiotensin II(AngII), whereas ACE2 hydrolyzes AngII into a series of vasodilators. There were few reports illustrated the expression of AngII in COVID-19. This study aimed to demonstrate the expression of angiotensin II in COVID-19 and how it correlated to the disease.

**Methods:** We enrolled 55 patients with COVID-19 admitted to renmin Hospital of Wuhan University from January 21st to February 21st, 2020. Demographic data were collected upon admission. COVID-19 nuclear acid, plasma AngII, Renin and aldosterone in the lying position without sodium restriction, and other laboratory indicators were together measured by the laboratory department of our hospital.

**Findings:** Of the 55 patients with COVID-19, 34(61.8%) had an increased level of AngII. The severity of COVID-19 and male is positively related with the level of AngII. The level of blood lymphocyte, PCT, ALT, and AST were remarkably severe with those of normal level of AngII (P < 0.05). CD4/CD8 cells ratio was significantly higher whereas CD3+CD8+ cells amount, CD3+CD8+ cells proportion, CD56+CD16+CD3- cells amount and CD19+CD3- cells amount were considerably lower than those of normal level of AngII (P < 0.05). Abnormal rates of blood lymphocyte and PCT were significantly higher in Patients with elevated AngII level. The results of binary logistic regression analysis showed that the severity of COVID-19 (OR=4.123) and CD4/CD8 ratio(OR=4.050) were the co-directional impact factor while female(OR=0.146) was inverse impact factor of elevated AngII level.

**Interpretation:** High rate of increased level of AngII was detected in COVID-19 patients. Patients with elevated AngII level were more likely to be critically ill with COVID-19. Considering the gender differences in ACE2 expression and no gender differences in angiotensin expression, the gender differences in AngII level might indicate less loss of ACE2 in female patients. Elevated AngII level was correlated with CD4/CD8 ratio, suggesting it might involve in immune disorder.

## Introduction

2019 Novel coronavirus disease(COVID-19) is rampant in China since December 2019 and spread worldwide gradually^1–5^. Up to March 17th, a total of 179111 confirmed cases and 7,426 dead cases were reported worldwide according to WHO updates^5^. Angiotensin-converting enzyme 2 (ACE2) is identified as an important functional receptor for SARS-Cov-2^6–7^. The host receptor ACE2 degrades after binding to SARS-Cov-2, leading to ACE2 loss and prompting the target oragns injury^6^. ACE2 and ACE are homologues with opposite functions in the renin– angiotensin system^8–9^. ACE converts angiotensin I into a vital vasoactive peptide called angiotensin II(AngII), whereas ACE2 hydrolyzes AngII into a series of vasodilators. Theoretically, the loss of ACE2 may reduce degradation of AngII and cause vasoconstriction and oxidative stress. Recently, a small sample study found that plasma angiotensin II levels were significantly increased and linearly associated to viral load and lung injury in COVID-19^10^.However, the sample size was too small to observe the exact relationship with the disease. Therefore, this study aimed to demonstrate the expression of angiotensin II in COVID-19 and how it correlated to the disease.

## Methods

### Study Design and Participants

This was a single center, retrospectively and observational analysis. We enrolled 55 patients with COVID-19 admitted to renmin Hospital of Wuhan University from January 21st to February 21st, 2020. All patients hadn’t taken any angiotensin-converting enzyme (ACE) inhibitors, angiotensin II type 1 receptor (AT1R) blockers, and diuretics two weeks before and during hospitalization. The diagnosis of hypertension and diabetes were based on 2018 ESC/ESH Guidelines for the management of arterial hypertension and 2019 ESC Guidelines on diabetes, pre-diabetes, and cardiovascular diseases developed incollaboration with the EASD.COVID-19 was diagnosed based on the Diagnosis and Treatment Scheme for New Coronavirus Pneumonia ( Pilot Edition 5, Revised version) published by the National Health Commission of China^11^. A confirmed case with COVID-19 was defined as a positive result to real-time reverse-transcriptase polymerase-chain-reaction (RT-PCR) assay for nasal and throat swab specimens. All patients had imaging pneumonia. Critically ill COVID-19 was defined as meeting either one of the flowing criteria: 1) Respiratory distress with respiratory rate more than 30 times/min; 2) Oxygen saturation ≤93% in resting state; 3) PaO2/FiO2 ≤300mmHg (1mmHg=0.133kPa).4) Respiratory failure requires mechanical ventilation; 5) Shock; 6) Combining other organ failures requires ICU monitoring and treatment. The study was approved by the ethics committee of renmin Hospital of Wuhan University(Application ID:[WDRY2020-K114]).

### Clinical and laboratory Data Collection

Demographic data including age, gender, and previous medical history were collected. Laboratory assessments consisting of plasma AngII, renin and aldosterone in the lying position without sodium restriction, complete blood count, blood chemistry, coagulation test, liver and renal function, electrolytes, C-reactive protein, procalcitonin, lactate dehydrogenase and creatine kinase were tested by the laboratory department.

### Statistical Analysis

All statistical analyses were performed by SPSS for mac software, version 23.0. Continuous variables were presented as the means and standard deviations or medians and interquartile ranges (IQR) as appropriate. Categorical variables were summarized as the counts and percentages in each category. Independent-Samples T test or the Mann-Whitney U test were applied to continuous variables, chi-square tests and Fisher’s exact tests were used for categorical variables as appropriate. Predictors of ANG II anomaly were analyzed by logistic regression. A value of p < 0.05 was considered statistically significant.

## Results

By Feb 21st, 2020, 55 confirmed cases of COVID-19 were included in this study. All of them had data on plasma AngII, renin and aldosterone in the lying position without sodium restriction. 34(61.8%) cases had an increased level of AngII while most patients had normal levels of renin and aldosterone (showed in table 1). The critically ill patients had higher level of AngII than the non-critically ill patients (showed in table 2).

**Table 1.** The baseline value of RAS system in COVID-19 patients

| RAS system parameter | Normal Range | Total (N=55) | Abnormality, N(%) |
| --- | --- | --- | --- |
| AngII(pg/ml, IQR) | 25-129 | 134.2(117.8,155.3) | 34(61.8) |
| Renin(pg/ml, IQR) | 4-24 | 10.9(6.3,16.9) | 11 (20) |
| ALD(pg/ml, IQR) | 10-160 | 129.5(112,149.8) | 9(16.4) |
| AARR(IQR) | / | 12.3(7.6,19.2) | / |

To further analyze the demographic, clinical and laboratory characteristics of the patients with increased ANG II level, we divided the patients into the AngII increased group and the AngII normal group. No difference was seen in renin and aldosterone values between the two groups (showed in table 3). To our interest, as shown in Table 4, the patients with increased level of AngII were more severe than those with normal level of AngII [18(52.9%)vs5(23.8%), p=0.033]. Significant gender differences were found between the two groups. In addition, there was no significant difference in the history of hypertension and the use of vasoactive drugs such as norepinephrine and dopamine in the two groups.

**Table 2.** Comparison of RAS system between non-critically ill and critically ill patients with COVID-19

| RAS system | Normal Range | Total (N=55) | Non-critically ill group(n=32) | critically ill group(n=23) | P value |
| --- | --- | --- | --- | --- | --- |
| AngII(pg/ml, IQR) | 25-129 | 134.2(117.8,155.3) | 127.4(93.5,145.6) | 147.4(131.5,169.5) | 0.009* |
| Renin(pg/ml, IQR) | 4-24 | 10.9(6.3,16.9) | 10.2(5.8,16.2) | 14.7(6.5,19.7) | 0.232 |
| ALD(pg/ml, IQR) | 10-160 | 129.5(112,149.8) | 133.4(115.1,168.1) | 115.9(104.8,135.7) | 0.050 |
| AARR(IQR) | / | 12.3(7.6,19.2) | 14.1(10.1,21.5) | 9.1(6.2,18.7) | 0.028* |
| **Ang II: angiotensin II, ALD:aldosterone, AARR: aldosterone/ Renin** |  |  |  |  |  |

**Table 3.** Comparison of RAS system between COVID-19 patients grouped by Ang II level

| RAS system | Normal Range | Total (N=55) | Ang II increased group(n=34) | Ang II normal group(n=21) | P value |
| --- | --- | --- | --- | --- | --- |
| AngII(pg/ml, IQR) | 25-129 | 134.2(117.8,155.3) | 149.7(137.8,165.1) | 99.4(84.5,119.1) | 0.000 |
| Renin(pg/ml, IQR) | 4-24 | 10.9(6.3,16.9) | 11.1(6.0,19.5) | 10(6.5,16.9) | 0.842 |
| ALD(pg/ml, IQR) | 10-160 | 129.5(112,149.8) | 133.4(112.4,142.1) | 121.8(106.9,163.5) | 0.591 |
| AARR(IQR) | / | 12.3(7.6,19.2) | 12.3(7.0,19.9) | 12.4(9.3,19.5) | 0.665 |
| **Ang II: angiotensin II, ALD:aldosterone, AARR: aldosterone/ Renin** |  |  |  |  |  |

**Table 4.**
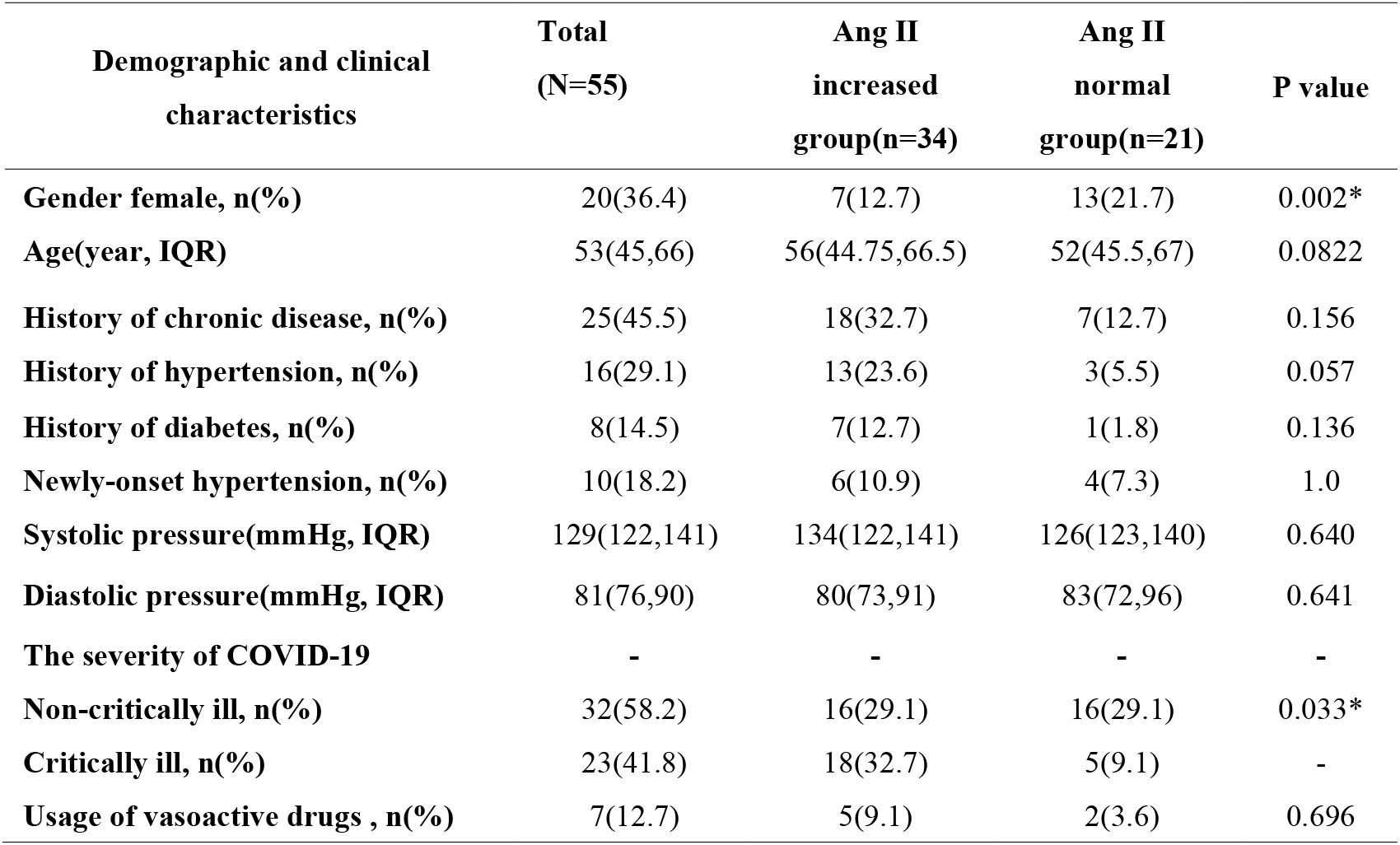
Comparison of Demographic and clinical characteristic between COVID-19 patients grouped by Ang II level

As presented in tables 5–6, there were statistical difference in the level of blood lymphocyte[0.66(0.36,1.03)vs1.02 (0.68, 1.42), p=0.021], PCT[0.07 (0.03, 0.120 vs0.03 (0.02, 0.07), p=0.007], CD4/CD8 cells ratio[2.35 (1.86, 3.22) vs1.55 (1.11, 2.54), p=0.015], ALT[27 (21, 44) vs19 (13, 34), p=0.03], AST[24.5(18.5,36)vs18(14,22.5),p=0.028], CD3+CD8+ cells [128(51,206)vs218(123,322),p=0.016], CD3+CD8+ cells proportion [20.1(14,25.6)vs25.4(20.9,35.7),p=0.011], CD56+CD16+CD3- cells [81(56,102)/111(66,171) p=0.031] between the AngII increased group and the AngII normal group. The rate of Lymphopenia [27(79.4%)vs11(52.4%), p=0.035] were remarkably higher in Patients with elevated AngII level.

**Table 5.** Comparison of RAS system between COVID-19 patients grouped by Ang II level

| **Laboratory assessments** | **Normal Range** | **Total (N=55)** | **Ang II increased group(n=34)** | **Ang II normal group(n=21)** | **P value** |
| --- | --- | --- | --- | --- | --- |
| **WBC($10^9$/L,IQR)** | 3.5-9.5 | 6.76(5.25,9.36) | 6.98(4.77,10.06) | 6.67(5.67,8.23) | 0.952 |
| **Ly ($10^9$/L,IQR)** | 1.1-3.2 | 0.74(0.49,1.19) | 0.66(0.36,1.03) | 1.02(0.68,1.42) | 0.021* |
| **HB(g/L,IQR)** | 130-175 | 128(114,142) | 127.5(112,142.8) | 128(114,140) | 0.959 |
| **PLT($10^9$/L,IQR)** | 125-350 | 198(142,263) | 189(122.3,263.8) | 238(171,274) | 0.225 |
| **CRP(mg/L,IQR)** | 0-8 | 7.73(2.88-26.36) | 11.2(3.67,44.99) | 7.03(1.53,21.20) | 0.253 |
| **PCT (ng/mL,IQR)** | 0-0.09 | 0.06(0.03,0.1) | 0.07(0.05,0.12) | 0.03(0.02,0.07) | 0.007* |
| **ALT(U/L,IQR)** | 9-50 | 26(18,38) | 27(21,44.3) | 19(13,34) | 0.030* |
| **AST (U/L,IQR)** | 15-40 | 21(16,32) | 24.5(18.5,36) | 18(14,22.5) | 0.028* |
| **ALB(g/L,IQR)** | 40-55 | 33.9(30.1,37.4) | 33.6(30.1,36.1) | 36(29.8,38.9) | 0.253 |
| **BUN(mmol/L,IQR)** | 3.2-8.0 | 4.9(3.0,7.5) | 4.95(3.1,8.38) | 4.3(2.85, 6.45) | 0.287 |
| **sCr(umol/L,IQR)** | Male:57-97<br>Female:41-81 | 60(48.2,67.4) | 61.3(54.6,70.3) | 55.2(45.7, 65.8) | 0.203 |
| **LDH(U/L,IQR)** | 120-250 | 214(179,354) | 244(188.3,400) | 201(176.5,258) | 0.057 |
| **CK(U/L,IQR)** | 50-310 | 39(26,57) | 38(26.8,59) | 40(25,59) | 0.972 |
| **Na(mmol/L,IQR)** | 137-147 | 135(132.1,138.7) | 134.9(131.6,137.5) | 136.3(134,138.9) | 0.194 |
| **K(mmol/L,IQR)** | 3.5-5.3 | 4.01(3.70,4.61) | 3.99(3.64,4.62) | 4.2(3.82,4.63) | 0.550 |
| **DD(mg/L,IQR)** | 0.01-0.55 | 1.96(0.69,4.92) | 3.46(0.82,6.56) | 1.2(0.63,30.2) | 0.121 |
| **APTT(s,IQR)** | 25-31.3 | 26.5(23.8,29.1) | 26.1(24.0,28.1) | 26.8(23.5,30.2) | 0.808 |
| **Urine protein positive, n(%)** | / | 7(12.7) | 6(10.9) | 1(1.8) | 0.232 |
| **Urine RBC positive, n(%)** | / | 3(5.35) | 2(3.64) | 1(1.8) | 1.0 |
| **Ly<1.1, n(%)** | / | 38(69.1) | 27(49.1) | 11(20) | 0.035* |
| **PCT>0.09, n(%)** | / | 15(27.3) | 12(21.8) | 3(5.5) | 0.123 |

**Abbreviation: Ly: Lymphocyte, HB: Hemoglobin, PLT: Platelet count, CRP: C-reactive protein, PCT: Procalcitonin, BUN: Blood urea nitrogen, sCr: serum creatinine, ALT :Aspartate aminotransferase, AST: Alanine aminotransferase, ALB: albumin, LDH: lactate dehydrogenase, CK: Creatinine kinase, DD: D-dimer, APTT: Activated partial thromboplastin time**

**Table 6.** Comparison of Lymphocyte classification between COVID-19 patients grouped by Ang II level

| **Lymphocyte marker** | **Normal Range** | **Total (N=46)** | **Ang II increased group(n=27)** | **Ang II normal group(n=19)** | **P value** |
| --- | --- | --- | --- | --- | --- |
| **CD3+ count (/uL,IQR)** | 1185-1901 | 561(274.3,776.3) | 439(178,741) | 669(448,841) | 0.073 |
| **CD3+ (% ,IQR)** | 64.19-75.77 | 68.9(60.75,77.93) | 69.1(55.5,82.1) | 68.7(66.4,77.6) | 0.510 |
| **CD3+CD4+ count (/uL,IQR)** | 561-1137 | 335.5(157.8,488.3) | 298(122,488) | 390(234,504) | 0.160 |
| **CD3+CD4+ (% ,IQR)** | 30.09-40.41 | 44.9(34.4,51.1) | 47.4(34.5,56.2) | 41.2(34,47.9) | 0.212 |
| **CD3+CD8+ count (/uL,IQR)** | 404-754 | 163(74.8,243.5) | 128(51,206) | 218(123,322) | 0.016* |
| **CD3+CD8+ (% ,IQR)** | 20.74-29.42 | 23.0(17.1,29.3) | 20.1(14,25.6) | 25.4(20.9,35.7) | 0.011* |
| **CD4+/CD8+** | 1.36-2.61 | 2.05(1.47,2.55) | 2.35(1.86,3.22) | 1.55(1.11,2.54) | 0.015* |
| **CD56+CD16+CD3-**<br>**count (/uL,IQR)** | 175-567 | 86.5(62.8,125.5) | 81(56,102) | 111(66,171) | 0.031* |
| **CD56+CD16+CD3-**<br>**(%,IQR)** | 10.04-19.78 | 13.75(7.95,20.73) | 12.9(7,24) | 13.8(8.3,18.9) | 0.832 |
| **CD19+CD3- count**<br>**(/uL,IQR)** | 180-324 | 91(55.8,155.8) | 73(46,131) | 134(59,170) | 0.072 |
| **CD19+CD3- (% ,IQR)** | 10.12-15.42 | 13.8(8.5,21.7) | 14.1(5.7,24.4) | 13.5(9.6,19.5) | 0.422 |

Furthermore, we evaluated the effect of various clinical and laboratory indicators on elevated AngII level with binary regression analysis. During the analysis, we applied AngII elevated or not as dependent variables, while applying the severity of COVID-19, gender, lymphocyte, PCT, CD4/CD8 cells ratio,CD3+CD8+ cells count, CD3+CD8+ cells proportion, CD56+CD16+CD3- cells count as independent variables, among these independent variables. The results showed that the severity of COVID-19 [OR=4.123, 95%CI(1.07-15.877), p=0.040] and CD4/CD8 ratio[OR=4.050, 95%CI(1.207-13.588), p=0.024]was the co-directional impact factor while female[OR=0.146,95%CI(0.035-0.603), p=0.008] were reverse impact factor of elevated AngII level(depicted in Figure 1).

**Figure 1:**
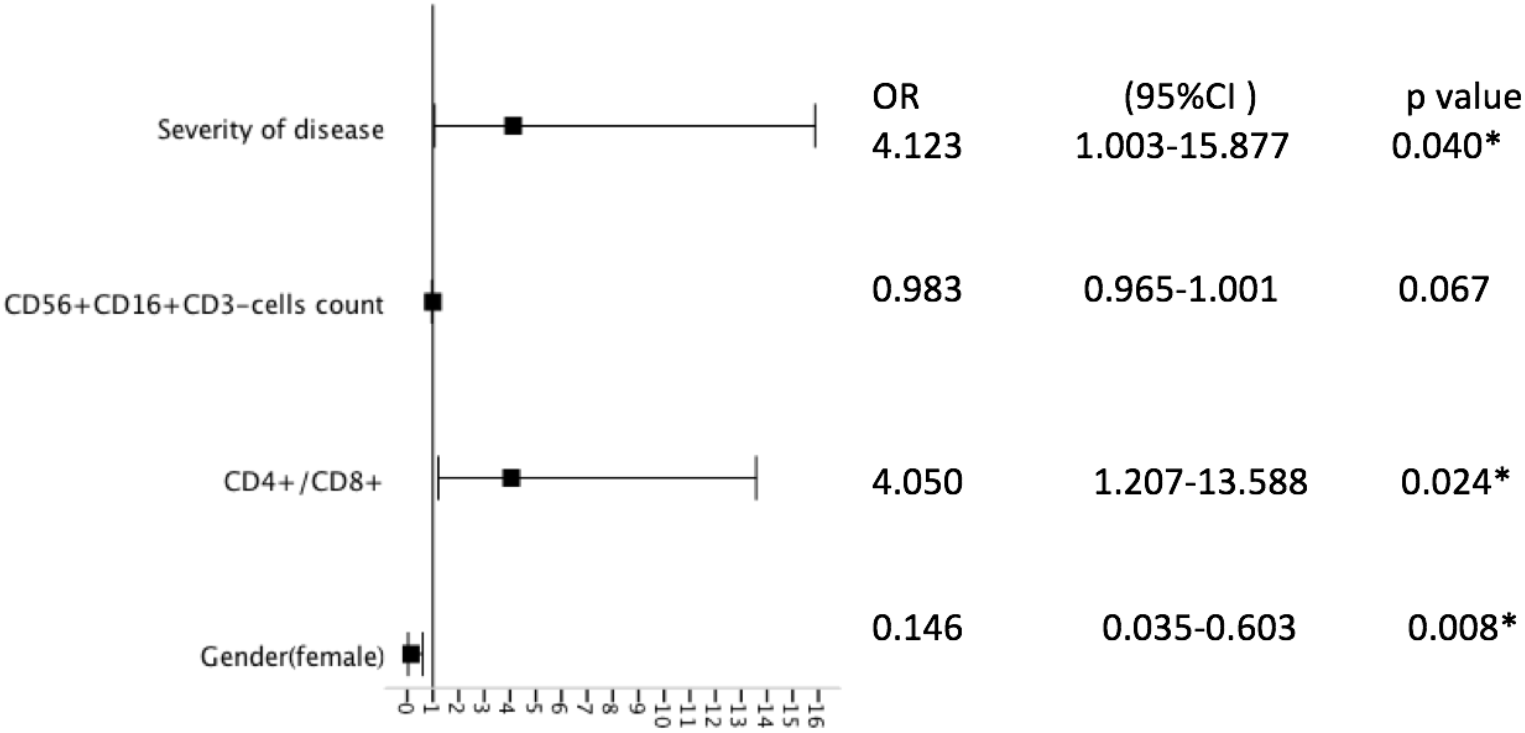
The effect of clinical and laboratory indicators on elevated Angll level with binary regression analvsis

## Discussion

As COVID-19 outbreak continues to spread globally, the newly discovered infectious disease may cause global public health crisis. It is reported that on February 28th, the World Health Organization raised the global risk of transmission and impact of COVID-19 to “very high” level^12^. Although some literatures have been published, as a new epidemic infectious disease, the epidemiological and clinical characteristics of COVID-19 are not well known. Human ACE2 is confirmed to be the receptor and a gateway for SARS-CoV-2^6^. ACE2 is a zinc metalloproteinase homologous to ACE, which can directly convert Ang II to Ang-(1-7), thus acting as a negative regulator of the renin-angiotensin system. Therefore, it could be hypothesized that a decrease in ACE2 caused by SARS-CoV-2 infection will reduce the degradation of Ang II, thereby causing an increase in Ang II^8–9^. A recently published small sample study not only confirmed this hypothesis, but also found that angiotensin II levels were linearly associated to viral load and lung injury in COVID-19^10^. This study reported a high rate of increased level of AngII in COVID-19 patients, which could be verified with the study above. There was no significant difference in the history of hypertension and the use of vasoactive drugs such as norepinephrine and dopamine in the two groups, which means that abnormality of AngII cannot be blamed to a history of hypertension and the use of vasoactive drugs.

This study showed a significant difference in the severity of COVID-19 in the elevated AngII group, and the severity of COVID-19 was a risk factor of increased AngII level. When grouped according to disease severity, AngII was remarkably higher in critically ill patients than those with mild disease. That implied that AngII level was closely related to disease severity.

Furthermore, this study found that the level of blood lymphocyte, CD3+CD8+ cells, CD56+CD16+CD3- cells, CD3+CD8+ cells proportion were dramatically lower and the CD4/CD8 cells ratio was higher in the elevated AngII group than the normal AngII group. In addition, CD4/CD8 cells ratio was a risk factor of increased AngII level. As we know, CD3+CD8+ cells, CD56+CD16+CD3- cells are killer cells that can recognize and eliminate virus-infected cells. This finding suggested that elevated AngII level may be associated with a reduction in killer cells. Accumulating evidences showed that CD8+ T cells are mediators of hypertension. hypertension in response to AngII treatment was reduced by ~50% in Cd8−/− mice^13–15^. These studies may provide clues to explain the finding that there was no statistically significant difference in the proportion of new hypertension in the two groups grouped by angiotensin levels. In addition, According to literature reports and our clinical observations, patients with COVID-19 often suffer from immune disorders and even immune storms^16^. What role AngII plays in immune disorders in COVID-19 needs further concern.

There is no gender difference in the mean baseline values for plasma Ang II among normal population^17^. However, this study revealed significant gender differences in the mean baseline values for plasma Ang II among COVID-19 patients. Since ACE2 gene is located on the X chromosome, and estrogen increases ACE2 expression, ACE2 expression is higher in female than male^18^.Considering the gender differences in ACE2 expression, the gender differences in AngII level might deduce less loss of ACE2 in female patients. However, the exact mechanism needs to be further explored.

In summary, high rate of increased level of AngII was detected in COVID-19 patients. AngII level seemed to relevant to the severity of the disease, gender differences and immune disorder. This study was a single-center, retrospective analysis of a small sample with many confounding factors. Therefore, the conclusions above need to be verified by strict prospective or experimental research.

## Data Availability

The raw/processed data required to reproduce these findings cannot be shared at this time as the data also forms part of an ongoing study.

## References

1. Wang C, Horby PW, Hayden FG, Gao GF. A novel coronavirus outbreak of global health concern [published correction appears in Lancet. 2020 Jan 29]. Lancet. 2020;395(10223):470–473. doi:10.1016/S0140-6736(20)30185-9.

2. Huang C, Wang Y, Li X, et al. Clinical features of patients infected with 2019 novel coronavirus in Wuhan, China [published correction appears in Lancet. 2020 Jan 30]. Lancet. 2020;395(10223):497–506. doi:10.1016/S0140-6736(20)30183-5.

3. Li Q, Guan X, Wu P, et al. Early Transmission Dynamics in Wuhan, China, of Novel Coronavirus-Infected Pneumonia [published online ahead of print, 2020 Jan 29]. N Engl J Med. 2020;10.1056/NEJMoa2001316. doi:10.1056/NEJMoa2001316.

4. China National Health Commission. March 17th, 2020, update on the novel coronavirus pneumonia outbreak. Beijing: National Health Commission of the People’s Republic of China, 2020. http://www.nhc.gov.cn/xcs/yqtb/202003/97b96f03fa3c4e8d8d0bf536271a10c0.shtml.(check at 3.18).

5. WHO. Coronavirus disease 2019 (COVID-19) Situation Report – 57. Data as reported by 10AM CET 17 March 2020. https://www.who.int/docs/default-source/coronaviruse/situation-reports/20200317-sitrep-57-covid19.pdf?sfvrsn=a26922f2_4. (check at 3.18)

6. Letko M, Marzi A, Munster V. Functional assessment of cell entry and receptor usage for SARS-CoV-2 and other lineage B betacoronaviruses [published online ahead of print, 2020 Feb 24]. Nat Microbiol. 2020;10.1038/s41564–020–0688-y. doi:10.1038/s41564-020-0688-y.

7. Wan Y, Shang J, Graham R, Baric RS, Li F. Receptor recognition by novel coronavirus from Wuhan: An analysis based on decade-long structural studies of SARS [published online ahead of print, 2020 Jan 29]. J Virol. 2020;JVI.00127–20. doi:10.1128/JVI.00127-20.

8. Santos RAS, Sampaio WO, Alzamora AC, et al. The ACE2/Angiotensin-(1–7)/MAS Axis of the Renin-Angiotensin System: Focus on Angiotensin-(1–7). Physiol Rev. 2018;98(1):505–553. doi:10.1152/physrev.00023.2016.

9. Arendse LB, Danser AHJ, Poglitsch M, et al. Novel Therapeutic Approaches Targeting the Renin-Angiotensin System and Associated Peptides in Hypertension and Heart Failure. Pharmacol Rev. 2019;71(4):539–570. doi:10.1124/pr.118.017129.

10. Liu Y, Yang Y, Zhang C, et al. Clinical and biochemical indexes from 2019-nCoV infected patients linked to viral loads and lung injury. Sci China Life Sci. 2020;63(3):364–374. doi:10.1007/s11427-020-1643-8.

11. China National Health Commission. the Diagnosis and Treatment Scheme for New Coronavirus Pneumonia ( Pilot Edition 5, Revised version). Beijing: National Health Commission of the People’s Republic of China, February 8th, 2020. http://www.nhc.gov.cn/yzygj/s7653p/202002/d4b895337e19445f8d728fcaf1e3e13a.shtml.

12. WHO. WHO Director-General’s opening remarks at the media briefing on COVID-19 - 28 February 2020. https://www.who.int/dg/speeches/detail/who-director-general-s-opening-remarks-at-the-media-briefing-on-covid19---28-february-2020.

13. Drummond GR, Vinh A, Guzik TJ, Sobey CG. Immune mechanisms of hypertension. Nat Rev Immunol. 2019;19(8):517–532. doi:10.1038/s41577-019-0160-5.

14. Trott, D. W. et al. Oligoclonal CD8+ T cells play a critical role in the development of hypertension. Hypertension 64, 1108–1115 (2014). This study implicates oligoclonal CD8+ T cells as the main T cell subset involved in promoting hypertension.

15. Liu, Y. et al. CD8+ T cells stimulate Na-Cl co-transporter NCC in distal convoluted tubules leading to salt-sensitive hypertension. Nat. Commun. 8, 14037 (2017).

16. China National Health Commission. March 3rd, 2020, the Diagnosis and Treatment Scheme for New Coronavirus Pneumonia (Pilot Edition 7). Beijing: National Health Commission of the People’s Republic of China, 2020.

17. Miller JA, Anacta LA, Cattran DC. Impact of gender on the renal response to angiotensin II. Kidney Int. 1999;55(1):278–285. doi:10.1046/j.1523-1755.1999.00260.x.

18. Te Riet L, van Esch JH, Roks AJ, van den Meiracker AH, Danser AH. Hypertension: renin-angiotensin aldosterone system alterations. Circ Res. 2015;116(6):960–975. doi:10.1161/CIRCRESAHA.116.303587.

